# Effective Contact Tracing for COVID-19: A Systematic Review

**DOI:** 10.1101/2020.07.23.20160234

**Authors:** Carl-Etienne Juneau, Anne-Sara Briand, Tomas Pueyo, Pablo Collazzo, Louise Potvin

## Abstract

**Background:** Contact tracing is commonly recommended to control outbreaks of COVID-19, but its effectiveness is unclear. This systematic review aimed to examine contact tracing effectiveness in the context of COVID-19.

**Methods:** Following PRISMA guidelines, MEDLINE, Embase, Global Health, and All EBM Reviews were searched using a range of terms related to contact tracing for COVID-19. Articles were included if they reported on the ability of contact tracing to slow or stop the spread of COVID-19 or on characteristics of effective tracing efforts. Two investigators screened all studies.

**Results:** A total of 32 articles were found. All were observational or modelling studies, so the quality of the evidence was low. Observational studies (n=14) all reported that contact tracing (alone or in combination with other interventions) was associated with better control of COVID-19. Results of modelling studies (n=18) depended on their assumptions. Under assumptions of prompt and thorough tracing with no further transmission, they found that contact tracing could stop an outbreak (e.g. by reducing the reproduction number from 2.2 to 0.57) or that it could reduce infections (e.g. by 24%-71% with a mobile tracing app). Under assumptions of slower, less efficient tracing, modelling studies suggested that tracing could slow, but not stop COVID-19.

**Conclusions:** Observational and modelling studies suggest that contact tracing is associated with better control of COVID-19. Its effectiveness likely depends on a number of factors, including how many and how fast contacts are traced and quarantined, and how effective quarantines are at preventing further transmission. A cautious interpretation suggests that to stop the spread of COVID-19, public health practitioners have 2-3 days from the time a new case develops symptoms to isolate the case and quarantine at least 80% of its contacts, and that once isolated, cases and contacts should infect zero new cases. Less efficient tracing may slow, but not stop, the spread of COVID-19. Inefficient tracing (with delays of 4-5+ days or less than 60% of contacts quarantined with no further transmission) may not contribute meaningfully to control of COVID-19.

**Funding:** LP holds the Canada Research Chair in Community Approaches and Health Inequalities (CRC 950-232541). This funding source had no role in the design, conduct, or reporting of the study.

**Competing interests:** CEJ has contractual agreements with the Centre intégré universitaire de santé et de services sociaux du Centre-Sud-de-l’Île-de-Montréal and is founder of Dr. Muscle and the COVID-19 Science Updates (https://covid1.substack.com/).

**Registration:** PROSPERO CRD42020198462

## INTRODUCTION

On March 11, 2020, the World Health Organization (WHO) characterized COVID-19 as a pandemic. A wide range of interventions have been proposed to control it, including contact tracing (Juneau et al. 2020). WHO guidelines for contact tracing state that “At least 80% of new cases [should] have their close contacts traced and in quarantine within 72 hours of case confirmation” (WHO, 2020). However, the WHO does not cite peer-reviewed evidence to support these guidelines, and some studies suggest that tracing needs to be more thorough and prompt to be effective (Ferretti et al. 2020; Hellewell et al. 2020). The US and European Centres for Disease Control and Prevention (CDC) also recommend contact tracing, but offer seemingly conflicting advice in the face of widespread transmission, when thousands of new contacts may need to be traced daily. Indeed, the US CDC states that “When a jurisdiction does not have the capacity to investigate a majority of its new COVID-19 cases, [it] should consider suspending or scaling down contact tracing” (US CDC, 2020). On the other hand, the European CDC advises that “Contact tracing should still be considered in areas of more widespread transmission, wherever possible, and in conjunction with physical distancing measures” (ECDC, 2020a). The ECDC also writes that “With increasing numbers of cases […] public health staff can be supplemented by people who do not have public health backgrounds” and that “With increased numbers of staff, the contact tracing operations require a greater degree of coordination” (ECDC, 2020b). This apparent contradiction and lack of evidence in official recommendations may leave contact tracers wondering which guidelines to follow. Would a large (and costly) contact tracing operation with new staff track enough contacts quickly enough to be effective? Case in point: the UK spent ten billion pounds on its test and trace programme, which may not have been effective (Iacobucci, 2020). How then might contact tracers be maximally effective? In carrying out this systematic review, we aimed to (1) examine contact tracing effectiveness and (2) identify characteristics of effective tracing efforts. Drawing from the evidence, we also aimed to (3) list and present targets for all key steps of the contact tracing process (such as how many and how fast contacts should be traced), and (4) propose evidence-based guidelines for effective tracing in the context of COVID-19. In the end, we hope that this systematic review will have practical relevance for public health practitioners looking to establish, operate, or oversee effective contact tracing efforts.

## METHODS

We prepared this systematic review in accordance with PRISMA guidelines (Moher et al. 2009) and registered its protocol with PROSPERO (CRD42020198462).

### Eligibility criteria

All studies evaluating the effectiveness of contact tracing efforts in the community (alone or in combination with other interventions) were included. Effectiveness was defined as stopping or slowing the spread of COVID-19 in usual conditions, with outcomes of interest including reproduction numbers and other measures of spread. Randomized trials, observational studies, and modelling studies were included. All methods of contact investigation were considered, including tracing by telephone or via mobile apps. Articles in English, French, Spanish, and Portuguese from all countries were included. Studies were included only if they focused on SARS-CoV-2, as this virus poses unprecedented challenges, notably in terms of spread and transmission (including asymptomatic and presymptomatic transmission). Abstracts, letters, protocols, preprints, and other unreviewed research were excluded, as well as studies limited to hospitals, nursing homes, prisons, and other enclosed spaces where transmission dynamics may not reflect that of the community. Reviews were also excluded, but their reference lists were checked for additional studies.

### Search strategy

MEDLINE (1946 - 2020 July 8), Embase (1974 - 2020 July 10), Global Health (1973 - 2020 Week 26), and All EBM Reviews (2005 - 2020 July 10) were searched using the terms “COVID-19” OR “coronavirus disease 2019” OR “SARS-CoV-2” OR “severe acute respiratory syndrome coronavirus 2” OR “2019-NCoV” OR “2019 novel coronavirus” AND “contact tracing” OR“contact-tracing” OR “tracing contact*” OR “contact follow-up” OR “case detection*” OR “contact investigation*” OR “epidemic investigation*” with no language or date restrictions. To find additional articles, we also reviewed reference lists, used the “related articles” and “cited by” functions in Google Scholar and PubMed, searched our own files, and consulted with colleagues.

### Data extraction and synthesis

Two investigators screened all studies and solved discrepancies by mutual agreement. Characteristics of studies were recorded in a spreadsheet, including: first author, publication date, study design, population, characteristics of contact tracing efforts, and main findings. Meta-analysis was not feasible due to substantial differences in study designs, outcomes, and effect measures. We did not assess risk of bias.

## RESULTS

### Result of the search

A total of 544 papers were found (results in the Supplement). Removing duplicates left 343. We retained 158 based on title, 64 based on abstract, and 27 based on full text. We found one additional study via reference lists and four more via the “cited by” and “similar articles” functions of PubMed and Google Scholar (eFigure in the Supplement). Therefore, this systematic review includes 32 studies (Table 1). All were observational or modelling studies, so the quality of the evidence was low. Results were generally consistent: 14 out of 14 observational studies (100%) and 16 out of 18 (89%) modelling studies reported that contact tracing (alone or in combination with other interventions) was associated with better control of COVID-19. However, we cannot rule out a potential publication bias, whereby results of ineffective tracing efforts would not be published.

### Observational studies

Observational studies (n=14) examined the association between contact tracing (in combination with other interventions) and transmission of COVID-19 in China (Bi et al.2020), Hong Kong (Cowling et al. 2020; Lam et al. 2020; Wong et al. 2020), Taiwan (Chen et al. 2020), Singapore (Ng et al. 2020), South Korea (Choi et al. 2020; Choi et al. 2020b), Vietnam (Dinh et al. 2020), France (Bernard Stoecklin et al. 2020), the US (Burke et al. 2020), in African countries (Nachega et al. 2020), and in international comparisons (Wilasang et al. 2020; Davalgi et al. 2020). All reported at least some benefit of contact tracing, although the independent contribution of tracing (vs. other interventions) could not be assessed.

### Modelling studies of contact tracing as part of a larger set of interventions

A number of modelling studies (n=11) examined contact tracing alone or in combination with other interventions to control outbreaks of COVID-19. In 5 studies, the combination of non-pharmaceutical interventions (NPIs) was shown to be effective in China to control the first outbreak (Zu et al. 2020; Maier and Brockmann, 2020; Tang et al. 2020a; Tang et al. 2020b; Lai et al. 2020). These interventions included case isolation, contact tracing, travel restrictions, social distancing, and hand washing, among others. Without any measures, there could have been a 67-fold increase in the number of COVID-19 cases according to Lai et al. (2020). However, in terms of contact tracing, one study showed that the number of contact traced in Wuhan as of January 22nd 2020 was insufficient compared to the population size and probably had limited impact on the epidemic control at first (Tang et al. 2020a).

In 5 other studies, the intertwined nature of diagnostic rate (mean time from onset of symptom to case diagnosis) and quarantine rate (proportion of contacts traced and quarantined) was clearly underlined (Goscé et al. 2020; Giordano et al. 2020; Kucharski et al. 2020; Tang et al. 2020c; Wu et al. 2020). For example, one study showed that diagnosis rate would have only minimal impact unless it is combined with improved quarantine efforts (Tang et al. 2020c). Similarly, Wu et al. (2020) showed that the reproduction number could be rapidly reduced under 1 with a combination of rapid testing and effective quarantine. This is in accord with Giordano et al. (2020)’s model of the pandemic in Italy, showing that the peak of the pandemic could have been reached sooner and the number of deaths lessened with increased population-wide testing and contact tracing. Accordingly, Kucharski et al. (2020) showed that combined case isolation and contact tracing was more effective in diminishing the reproduction number (to 0.94) than mass testing alone (2.5) compared to no measure (2.6). Adding physical distancing and app-based tracing further decreased the reproduction number in this model (0.87). Similarly, a study based on data from London estimated that adding different measures such as weekly universal testing, case isolation, contact tracing and facemask use to lockdown could reduce cumulative deaths by 48% compared to lockdown alone (Goscé et al. 2020). Hence, the combination of multiple measures may be key in controlling COVID-19.

Concerning contact tracing more specifically, the quarantine rate appeared to be crucial in these articles. Indeed, many showed the possibility of decreasing the number of cumulative cases of COVID-19 with an increased quarantine rate (Tang et al. 2020a; Tang et al. 2020c; Wu et al. 2020; Zu et al. 2020). In one model (Tang et al. 2020c), it was estimated that a rate of only 0.6 (60% of contacts traced and isolated) could lead to a Rc below 1 and that the rate could be even less with some degree of social distancing maintained. Yet, this finding was not generalizable to all the pandemic phases, with unrealistic values of quarantine rate needed to avoid a rebound in the de-escalation phase where schools were reopened. In another study, based on China’s data, increasing the quarantine rate by 10 times led to a reduction of the peak number of infected individuals by 87% (Tang et al. 2020a). However, it was specified that such high values of quarantine would require important public health resources to control an outbreak. Correspondingly, Kucharski et al. (2020) estimated that a high level of tracing was required to ensure a reproduction number below 1. Lastly, a model looking at transmission dynamics in the state of New York and the US also found that only a small decrease in the cumulative cases and deaths would be derived from a high level of contact traced (Ngonghala et al. 2020).

### Modelling studies of contact tracing efforts

A small number of modelling studies (n=5) examined variations in contact tracing efforts and their impact on effectiveness. Broadly, they found that tracing effectiveness depended on how many individuals in the community are infected; how fast new cases are tested and isolated; how many of their contacts are traced and quarantined; how fast those contacts are quarantined; and how effective quarantines and isolations are at preventing further transmission. Keeling et al. (2020) estimated that under “optimistic assumptions”, with each step executed flawlessly, the basic reproduction number (R0) of an epidemic could be reduced from 3 to 0.18. When the percentage of contacts traced decreased from 100% to just over 71%, R0 decreased to just below 1, suggesting that 71% may be the lowest target to aim for. Similarly, Ferretti et al. (2020) modelled an epidemic with an R0 of 2.0 and assumed perfectly successful quarantine of contacts upon tracing, and perfect prevention of transmission from cases upon isolation. Even under these assumptions, a 3-day delay from initiation of symptoms to case isolation and quarantine of contacts made these interventions ineffective at controlling the epidemic. For a 2-day delay, about 80% of cases needed to be isolated and 80% of their contacts traced and quarantined to control the epidemic. For a 1-day delay, about 70% case isolation and 70% contact tracing and quarantine was needed. These results suggest that delays have a substantial effect, and that even under optimistic assumptions, they should be lower than 3 days. Similarly, Kretzschmar et al. (2020) found that with a testing delay of 3 days or longer, even the most efficient tracing could not bring the reproduction number below 1. In that study, in the most optimistic scenario, testing and tracing delays were set to 0, and 100% of contacts were traced. Assuming around 40% of transmissions occurring before symptom onset and no further transmission upon quarantine, one model predicted that tracing could reduce the reproduction number from 1.2 to 0.8. A testing delay of more than 1 day required the tracing delay to be at most 1 day or at least 80% of contacts to be traced to keep the reproduction number below 1, illustrating that as delays increase, more contacts need to be traced and quarantined for a programme to be effective. Under more realistic assumptions, Hellewell et al. (2020) modelled a wide range of scenarios, with varying number of initial cases (5, 20, 40), R0 (1.5, 2.5, 3.5), delays (3.43 or 8.09 days), and contacts traced (0%, 20%, 40%, 60%, 80%, 100%). In all scenarios, isolation was assumed to be 100% effective at preventing further transmission. The number of initial cases had a large effect on the probability of achieving control. With an R0 of 2.5 and a 3.43-day delay from symptom onset to isolation, with five initial cases, there was a greater than 50% chance of achieving control in 3 months, even when only 20% of contacts were traced. However, with 40 initial cases, tracing 20% of contacts lead to fewer than 5% of simulations controlled; 80% contact tracing was necessary to control just under 80% of simulations within 3 months, illustrating the need for swift action. Likewise, as in other modelling studies, the delay from symptom onset to isolation had a substantial effect. With an R0 of 2.5, 20 initial cases, and 80% of contacts traced, the probability of achieving control decreased from 89% to 31% when the delay from symptom onset to isolation was increased from 3.43 to 8.09 days. These results suggest that to be effective, contact tracing should start as soon as cases appear in the community, at least 80% of contacts should be traced, and delays from symptom onset to isolation should not exceed 3.43 days. Even so, the authors concluded that in most plausible scenarios, tracing alone was insufficient to control an outbreak within 3 months. In perhaps the most realistic modelling study, Peak et al. (2020) examined two combinations of parameters. In the first combination, (the “high-feasibility setting”): (1) 90% of contacts were traced; (2) the delay was 0.5 days; (3) quarantine reduced infectiousness by 75%; (4) isolation of cases reduced infectiousness by 90%. In this setting, the epidemic was controlled. Indeed, R0 was reduced from 2.2 to a median of 0.49 (with the longer serial interval of 7.5 days) or 0.57 (with the shorter serial interval of 4.8 days). According to the authors, this represented the upper bounds of efficiency expected of contact tracing efforts. In the second combination (the “low-feasibility setting”): (1) 50% of contacts were traced; (2) the delay was 2 days; (3) quarantine reduced infectiousness by 25%; (4) isolation of cases reduced infectiousness by 50%. In this setting, the epidemic was not controlled in any scenario, even when R0 was as low as 1.5. These results suggest that inefficient contact tracing may not contribute meaningfully to control of COVID-19. Overall, 5 out of 5 studies (100%) that modelled contact tracing efforts under optimistic assumptions suggested that contact tracing could stop an outbreak of COVID-19. On the other hand, 4 out of 4 studies (100%) that modelled less optimistic assumptions suggested that contact tracing alone could slow, but not stop an outbreak of COVID-19.

In addition, two modelling studies did not examine details of contact tracing efforts, but instead focused only on the adoption of a mobile phone tracing app (Yasaka et al. 2020; Currie et al. 2020). Yasaka et al. (2020) estimated that app-based contact tracing could reduce the proportion of population infected (about 80% infected with no tracing, about 55% with 25% adoption of the tracing app, about 45% with 50% adoption, and about 35% with 75% adoption). Likewise, Currie et al. (2020) found that in the baseline scenario representative of the ongoing situation in Australia, COVID-19 case count was reduced by an estimated 24% with 27% adoption of the app (actual adoption as of May 20, 2020), 32% with 40% adoption, 55% with 61% adoption, and 71% with 80% adoption). These studies did not model parameters such as delays from symptom onset to isolation and quarantine effectiveness, thus making optimistic assumptions which could explain the high reductions estimated.

## DISCUSSION

This systematic review aimed to examine contact tracing effectiveness and to identify characteristics of effective tracing programmes. We found that 15 out of 15 observational studies (100%) and 16 out of 18 (89%) modelling studies reported that contact tracing (alone or in combination with other interventions) was associated with better control of COVID-19. This conclusion is supported by a number of preprints and other unpublished work (Aleta et al. 2020; James et al. 2020; Worden et al. 2020; BC Ministry of Health, 2020).

Our results are in line with those of other reviews. A recent Cochrane review of quarantine (alone or in combination with other public health measures) found that modelling studies consistently reported a benefit of quarantine to control COVID^-^19, and that early implementation of quarantine and combining quarantine with other public health measures were important to ensure effectiveness (Nussbaumer-Streit et al. 2020). Likewise, a narrative review of contact tracing in patients infected with SARS-CoV-2 concluded that this “classic” strategy could be applied, but that it should be accelerated (Bellmunt et al. 2020).

Evidence-based guidelines for effective tracing in the context of COVID-19

Drawing from the evidence, this systematic review also aimed to list and present targets for all relevant dimensions of contact tracing, and to propose evidence-based guidelines. To do so, we can break down the steps of the contact tracing process as follows: (1) a new case develops symptoms; (2) the case is tested; (3) if positive, the case is isolated; (4) its contacts are traced; (5) contacts are notified and quarantined; (6) contacts are monitored during quarantine to prevent further transmission. Therefore, contact tracing effectiveness depends on how many individuals in the community are infected; how fast new cases are tested and isolated; how many of their contacts are traced and quarantined; how fast those contacts are quarantined; and how effective isolations and quarantines are at preventing further transmission. We propose guidelines for each of these dimensions in Table 2, along with supporting evidence. This evidence suggests that the current WHO guideline of tracing contacts within 3 days of cases being confirmed may at best slow, but not stop, the spread of COVID-19 (WHO, 2020).

**Table 2.** Evidence-based guidelines for effective contact tracing of COVID-19

|  | **Guidelines** |  |  |  |
| --- | --- | --- | --- | --- |
| **Dimension of contact tracing programme** | **Highly efficient**<br>**(can stop the spread)** | **Less efficient**<br>**(can slow the spread)** | **Inefficient**<br>**(may not contribute meaningfully)** | **Evidence base** |
| How many individuals in the community are infected | 5 or less | 20 or less | 40+ | Hellewell et al. 2020 |
| How fast new cases are tested and isolated* | 1 day or less | 1-2 days | 2+ days | Ferretti et al. 2020;<br>Kretzschmar et al. 2020 |
| How many of their contacts are traced and quarantined | 80% or more | 71-80% | Less than 71% | Ferretti et al. 2020;<br>Hellewell et al. 2020;<br>Keeling et al. 2020;<br>Kretzschmar et al. 2020 |
| How fast those contacts are quarantined* | 1-2 days | 2-3 days | 3+ days | Ferretti et al. 2020;<br>Kretzschmar et al. 2020 |
| How effective isolations and quarantines are at preventing transmission | 100% | 90-100% | Less than 75% | Ferretti et al. 2020;<br>Hellewell et al. 2020;<br>Peak et al. 2020;<br>Kretzschmar et al. 2020 |

\*Delays in isolation of cases and quarantine of contacts are combined in most studies. We break them down here as distinct steps of the contact tracing process. For highly efficient tracing, they should not exceed 2-3 days.

Contact tracing begins with a new case. The case develops symptoms, or is suspected (e.g. by being in close contact with another, confirmed case). The case can transmit the virus to others while presymptomatic or asymptomatic (Furukawa et al. 2020), therefore speed at this step is crucial (Keeling et al. 2020; Ferretti et al. 2020; Kretzschmar et al. 2020; Hellewell et al. 2020; Peak et al. 2020). The case should be tested and its results communicated in the shortest possible time. If new cases are not tested, wait to be tested, or wait for test results to be communicated, contact tracing becomes less effective. Thus testing availability and efficiency are important at this step. Testing should be widely available, and the public could be reminded to seek testing at the earliest signs of symptoms. Likewise, systemic delays in communicating test results should be examined and minimised. In the UK, anecdotal evidence suggests that test results were communicated in 2-3 days (Vize, 2020), which is likely to have undermined tracing efforts considerably. In the US, one report of drive-through testing found that when outsourced, tests (n=476) were turned around in a median of 9.21 days (Lindholm et al. 2020). Contact tracing may not be effective with such delays.

Once a case is confirmed, its contacts must be traced. Tracing contacts manually over the telephone has a number of limitations. First, it is likely that not all contacts will be traced. Cases may not disclose, remember, or have contact information for all contacts. For example, a pilot programme in Sheffield, UK, found that two thirds of people contacted through tracing did not fully cooperate (Mahase, 2020). Second, during a peak of COVID-19, thousands of new contacts may need to be traced daily. This poses substantial operational challenges. Large contact tracing efforts with new staff are costly, and may not be able to maintain the level of effectiveness of smaller programmes with experienced staff. For example, UK’s test and trace programme cost ten billion pounds and may not have been effective (Iacobucci, 2020). Mobile phone apps and other technologies can circumvent these shortcomings, but raise a number of ethical issues (Bradford et al. 2020; Austin et al. 2020) and pose technological challenges (Ivers and Weitzner, 2020). Moreover, early evidence suggests that contact tracing apps, despite wide encouragement, have limited adoption (Sim and Lim, 2020; Johnson, 2020). Thus, they may only contribute to slowing (but not stopping) spread of COVID-19. Other key steps of the contact tracing process are isolations and quarantines. Ineffective isolations and quarantines may compromise otherwise flawless tracing efforts. Four of the five modelling studies we have reviewed assumed perfect prevention of transmission at these steps (Keeling et al. 2020; Ferretti et al. 2020; Kretzschmar et al. 2020; Hellewell et al. 2020). This may be difficult to achieve in practice, even if cases and contacts never leave the home. Indeed, Bi et al. (2020) found that household secondary attack rate was 11.2% (95% CI 9.1–13.8) in Shenzhen, China, despite contact tracing being in place. Similarly, Park et al. (2020) found that 11.8% of household contacts had COVID-19 in South Korea. Moreover, Wu and McGoogan (2020) report that in 20 Chinese provinces outside of Hubei, a total of 1183 case clusters were found, of which 64% were within familial households. Thus it may be more effective to isolate and quarantine outside the home, in hotels or central locations, especially considering many homes (1 in 5 in the US) will lack sufficient space to comply with recommendations (Sehgal et al. 2020). If home isolation and quarantine are used nonetheless, information could be provided to new cases and contacts to reduce household transmission. Li et al. (2020) followed 105 index patients and 392 household contacts in Wuhan, China. They found 14 cases had isolated by themselves at home immediately after the onset of symptoms— with masks, dining separately, and residing alone. The results showed no infected contacts in the households of these index cases. A final consideration for this step of the process is financial and social support. Contacts may need support for lost wages, daily activities carried out outside the home (e.g. groceries), or questions related to their health (e.g. telephone helpline).

### Strengths and limitations

This study has a number of strengths. To our knowledge, this is the first study to systematically review the effectiveness of contact tracing efforts, and to propose evidence-based guidelines for tracing in the context of COVID-19. While an earlier review of quarantine found only modelling studies for COVID-19 (Nussbaumer-Streit et al. 2020), we found both observational and modelling studies. Therefore, both the overall association of contact tracing with transmission (in observational studies) and the influence of variations in dimensions of contact tracing efforts on their effectiveness (in modelling studies) could be examined. However, the nature of this body of evidence is also a limitation, as its quality is low, limiting the strength of our conclusions. In addition, we did not assess risk of bias, examine publication bias, or rate the certainty of evidence using GRADE guidelines. We plan to update this paper to address these limitations shortly. Moreover, most observational studies reported results of contact tracing efforts in combination with other interventions, so their individual contribution could only be estimated from modelling studies. Another limitation is the rapidly evolving nature of the pandemic (as of July 2020) and related contact tracing policies and regulations, as well as public perceptions, which are all likely to influence future tracing efforts. A final limitation is we did not discuss implementation—Rajan et al. (2020) describe specific challenges and solutions.

## Conclusions

Contact tracing effectiveness depends on how many individuals in the community are infected; how fast new cases are tested and isolated; how many of their contacts are traced and quarantined; how fast those contacts are quarantined; and how effective quarantines and isolations are at preventing further transmission. A cautious interpretation of the evidence suggests that to stop the spread of COVID-19 with contact tracing, public health practitioners have 2-3 days from the time a new case develops symptoms, to isolate the case and quarantine at least 80% of its contacts, and that once isolated, cases and contacts should infect zero new cases. The evidence suggests that less efficient tracing may slow, but not stop, the spread of COVID-19, and that inefficient tracing (with delays of 4-5+ days and less than 60% of contacts quarantined with no further transmission) may not contribute meaningfully to control of COVID-19. Considering the potential human and economic costs of inefficient tracing, we hope that these guidelines can help public health practitioners establish and oversee effective contact tracing efforts. Future research may improve our understanding of their effectiveness by assessing emerging empirical evidence from ongoing efforts, best practices and policy responses, and differences in outcomes across jurisdictions with more or less efficient tracing.

## Data Availability

All data available upon request.

## SUPPLEMENT—RESULT OF THE SEARCH

| Step | Searches | Results |
| --- | --- | --- |
| 1 | COVID-19.mp. [mp=ti, ot, ab, nm, hw, kw, fx, kf, ox, px, rx, ui, an, sy, tx, sh, ct, tn, dm, mf, dv, dq, bt, id, cc] | 56118 |
| 2 | SARS-CoV-2.mp. [mp=ti, ot, ab, nm, hw, kw, fx, kf, ox, px, rx, ui, an, sy, tx, sh, ct, tn, dm, mf, dv, dq, bt, id, cc] | 17748 |
| 3 | 2019-NCoV.mp. [mp=ti, ot, ab, nm, hw, kw, fx, kf, ox, px, rx, ui, an, sy, tx, sh, ct, tn, dm, mf, dv, dq, bt, id, cc] | 1953 |
| 4 | coronavirus disease 2019.mp. [mp=ti, ot, ab, nm, hw, kw, fx, kf, ox, px, rx, ui, an, sy, tx, sh, ct, tn, dm, mf, dv, dq, bt, id, cc] | 29369 |
| 5 | severe acute respiratory syndrome coronavirus 2.mp. [mp=ti, ot, ab, nm, hw, kw, fx, kf, ox, px, rx, ui, an, sy, tx, sh, ct, tn, dm, mf, dv, dq, bt, id, cc] | 18947 |
| 6 | 2019 novel coronavirus.mp. [mp=ti, ot, ab, nm, hw, kw, fx, kf, ox, px, rx, ui, an, sy, tx, sh, ct, tn, dm, mf, dv, dq, bt, id, cc] | 1849 |
| 7 | 1 or 2 or 3 or 4 or 5 or 6 | 61984 |
| 8 | contact tracing.mp. [mp=ti, ot, ab, nm, hw, kw, fx, kf, ox, px, rx, ui, an, sy, tx, sh, ct, tn, dm, mf, dv, dq, bt, id, cc] | 9591 |
| 9 | contact-tracing.mp. [mp=ti, ot, ab, nm, hw, kw, fx, kf, ox, px, rx, ui, an, sy, tx, sh, ct, tn, dm, mf, dv, dq, bt, id, cc] | 9591 |
| 10 | tracing contact*.mp. [mp=ti, ot, ab, nm, hw, kw, fx, kf, ox, px, rx, ui, an, sy, tx, sh, ct, tn, dm, mf, dv, dq, bt, id, cc] | 79 |
| 11 | contact follow-up.mp. [mp=ti, ot, ab, nm, hw, kw, fx, kf, ox, px, rx, ui, an, sy, tx, sh, ct, tn, dm, mf, dv, dq, bt, id, cc] | 177 |
| 12 | case detection*.mp. [mp=ti, ot, ab, nm, hw, kw, fx, kf, ox, px, rx, ui, an, sy, tx, sh, ct, tn, dm, mf, dv, dq, bt, id, cc] | 8391 |
| 13 | contact investigation*.mp. [mp=ti, ot, ab, nm, hw, kw, fx, kf, ox, px, rx, ui, an, sy, tx, sh, ct, tn, dm, mf, dv, dq, bt, id, cc] | 2259 |
| 14 | epidemic investigation*.mp. [mp=ti, ot, ab, nm, hw, kw, fx, kf, ox, px, rx, ui, an, sy, tx, sh, ct, tn, dm, mf, dv, dq, bt, id, cc] | 267 |
| 15 | 8 or 9 or 10 or 11 or 12 or 13 or 14 | 19697 |
| 16 | 7 and 15 | 544 |
| 17 | remove duplicates from 16 | 343 |

## SUPPLEMENT

### eFigure—PRISMA Flow Diagram

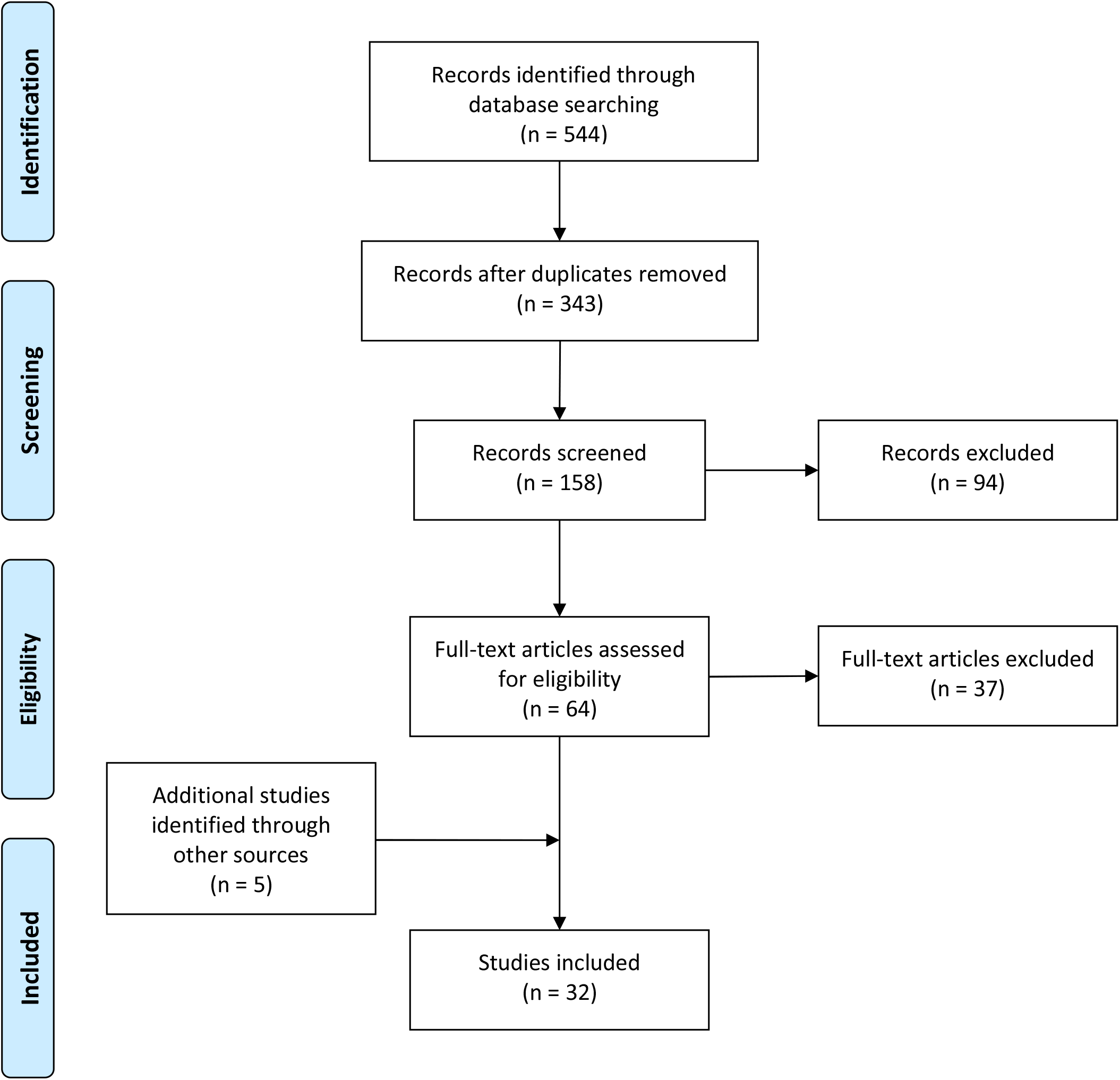

*From:* Moher D, Liberati A, Tetzlaff J, Altman DG, The PRISMA Group (2009). *P*referred *R*eporting *I*tems for *S*ystematic Reviews and *M*eta-*A*nalyses: The PRISMA Statement. PLoS Med 6(7): e1000097. doi:10.1371/journal.pmed1000097

**For more information, visit www.prisma-statement.org**.

